# Detection of risk factors of hypertension in people with sleep disorders using machine learning

**DOI:** 10.1101/2024.06.10.24308738

**Authors:** Wu Wenzhong, Liu Weixiang, Ye Junlan, Zhang Yuting, Qin Shan, Wang Xiaoqiu, Li Xiaojie, Xu Min, Lin Liyu, Liu Chengyong, Lian Xiaoyang, Yang Tao

## Abstract

**Background:** Sleep disorders are one of the etiologic factors in the development of hypertension, and the risk of hypertension in patients with sleep disorders is increasing due to the increase in the prevalence of sleep disorders in recent years. We hypothesized that machine learning could establish a prediction model for hypertension in patients with sleep disorders.

**Methods:** The data for patients diagnosed with sleep disorders were collected from two hospitals from 2019 to 2023, including medical history and biochemical indicators. After data processing, Logistic Regression, Decision Tree, Artificial Neural Network, Support Vector Machine, Naive Bayes, Adaptive Boosting, Random Forest, and Extreme Gradient Boosting (XGBoost) were selected for training. Five-fold cross-validation was used to evaluate the models, and accuracy, precision, recall, F1-score, and the area under the curve (AUC) were used to verify the discrimination and clinical practicability of the models. SHapley Additive exPlanation (SHAP) was used to explain the optimal model.

**Results:** The XGBoost model was superior to other models, with an accuracy of 0.7216, a precision of 0.7576, a recall of 0.7660, an F1-score of 0.7430, and an AUC of 0.844. The SHAP results showed that age, weight, white blood cells, creatine, uric acid, glycosylated hemoglobin, and platelets were the risk factors of hypertension in people with sleep disorders, while high density lipoprotein cholesterol was a protective factor.

**Conclusion:** Machine learning algorithms established a predictive risk model of hypertension in people with sleep disorders, which provides significant guidance to prevent and treat hypertension in people with sleep disorders.

## INTRODUCTION

People with sleep disorders typically experience prolonged sleep latency, trouble maintaining sleep, or inability to return to sleep [1]. Globally, approximately 25% of people report themselves as having a sleep disorder [2], and the prevalence of sleep disorders in China is as high as 39.1% [3]. Sleep disorders negatively affect the quality of life and health status of patients, including reduced cognitive performance, altered immunological function, and higher incidence of cardiovascular diseases, such as hypertension [4]. Studies have shown that the prevalence of hypertension is enhanced by 50% in the sleep-disordered population [5], and improving sleep conditions can assist in reducing blood pressure levels in patients [6]. Hence, it is clinically crucial to establish the risk factors for high blood pressure related to sleep disturbances.

Risk factors for the development of sleep disorder-induced hypertension are still being explored. Previous studies have suggested that age, sex, weight, and minimum arterial oxygen saturation are risk factors for the development of hypertension in sleep disorders, which were identified through simple correlation analyses of polysomnographic or demographic data [7–9]. However, these results are relatively one-sided and lack clinical applicability. Sleep disorders can lead to hypertension through complex mechanisms, such as activating the sympathetic nervous system [10] and triggering an inflammatory state [11], and clinical biochemical indexes are essential to reflect the status of sleep-disordered populations and to understand the pathogenesis. Risk factors screened from these indexes might have broader clinical applicability. With the advances in artificial intelligence technology, it is possible to identify the risk factors for sleep disorders leading to hypertension and build accurate and effective models for precise diagnosis and treatment using machine learning.

Machine learning methods are commonly used to learn patterns and trends in clinical data, which allows for the identification of high-risk indicators from numerous parameters [12]. To the best of our knowledge, there are few machine-learning-based models to predict hypertension from sleep disorders. Furthermore, diverse conditions can induce hypertension from sleep disorders;hence, existing models suffer from the drawbacks of difficulty in acquiring validated features associated with hypertension and reliance on a single modeling technique. This study proposed to find the correlation between common clinical indicators and the occurrence of hypertension in patients with sleep disorders using machine learning, so as to rapidly and effectively predict the risk of sleep disorder-induced hypertension.

## Methods

### Study population

The diagnostic codes of sleep disorders, including insomnia (G47.000x001) and sleep disorder (G47.900), were examined in the electronic medical systems of Jiangsu Provincial Hospital of Chinese Medicine and Jiangsu Provincial Government Hospital from October 2019 to October 2023. 7024 electronic medical records of patients with sleep disorders were collected, containing a total of 194 indicators, such as general information (sex, age, weight, and diagnosis), laboratory data (including routine blood indexes, hypersensitive C-reactive protein, blood biochemistry, routine urine tests, routine fecal tests, glycosylated hemoglobin, thyroid function, and coagulation function), and functional examinations (e.g., electrocardiogram, carotid artery color doppler ultrasound, chest computed tomography, cranial magnetic resonance (MR)). We included all the recorded indicators as far as possible to screen the risk factors for sleep disorders leading to hypertension. These data made this a retrospective study, which was approved by the Ethics Committee of Jiangsu Provincial Hospital of Chinese Medicine (Approved No. of Ethic Committee: 2021NL-202-02) and was registered as a clinical trial (Registration number: ChiCTR2200059161). All patients signed a consent form for testing at the time of admission, and all medical records and demographic information were completely anonymized before data analysis.

### Inclusion criteria

Sleep disorders: The patient was diagnosed as having a sleep disorder by the clinician according to the following inclusion criteria: (a) Difficulty initiating and/or maintaining sleep, defined as a sleep onset latency and/or wake after sleep onset greater than or equal to 30 min, and waking up earlier than desired, with a corresponding sleep time of less than or equal to 6.5 hours per night; or sleep disturbance (or associated daytime fatigue) causing significant distress or impairment in social, occupational, or other areas of functioning; (b) the presence of insomnia for at least 3 nights per week and for at least 3 months. This definition represents a combination of criteria from the American Academy of Sleep Medicine, the International Classification of Sleep Disorders, and the Diagnostic and Statistical Manual of Mental Disorders, along with quantitative cutoffs typically used in insomnia research.

Hypertension: The patient has a medical history of hypertension in the past and is currently taking oral antihypertensive drugs.

### Exclusion criteria

(a) Sleep disorders were not specifically described in the patient’s medical history; (b) Considering that the diagnostic criterion for patients with sleep disorders was 3 months, we doubled this and excluded hypertensive patients with a history of sleep disorders of less than 6 months.

### Framework

Having acquired the data on sleep disorders, a framework to analyze hypertension was proposed. As shown in Fig. 1, at first, data for sleep disorders were collected and preprocessed, and divided into a training set and a validation set. Then, multiple widely used machine learning algorithms including Logistic Regression (LR), Decision Tree (DT), Artificial Neural Network (ANN), Random Forest (RF), Support Vector Machine (SVM), Naive Bayes (NB), Extreme Gradient Boosting (XGBoost), and Adaptive Boosting (AdaBoost) were used to construct the predictor. Next, the final model was selected and interpreted based on model evaluation using K-Fold cross-validation. Eventually, it was concluded that age, weight, white blood cell count (WBC), creatinine (Cr), uric acid (UA), glycosylated hemoglobin (HbA1c), high-density lipoprotein cholesterol (HDLC), and platelets are the important characteristics of hypertension caused by sleep disorders.

**Fig. 1.**
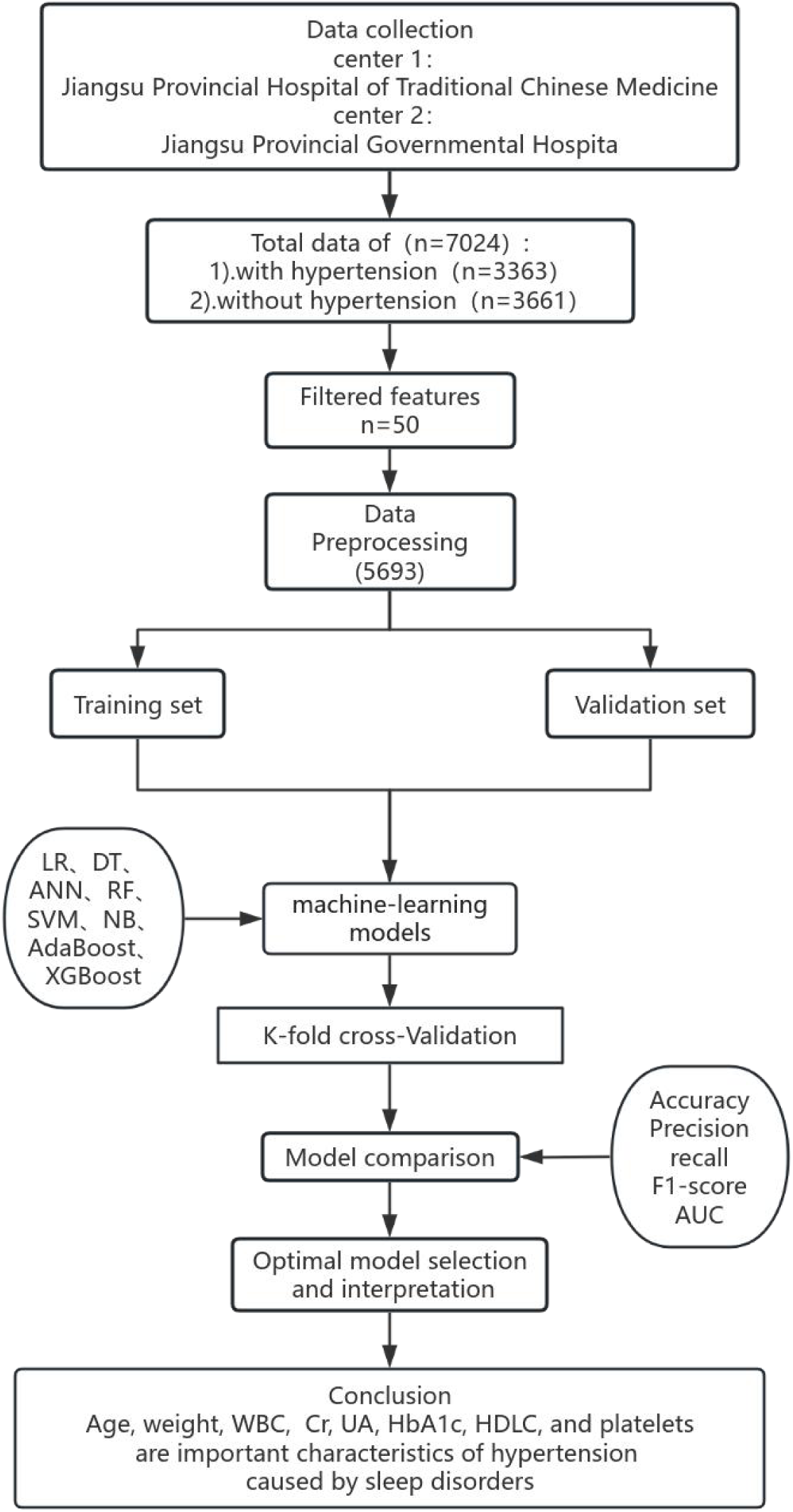
Research design to establish a machine learning model to predict hypertension after sleep disorder. LR, Logistic Regression; DT, Decision Tree; ANN, Artificial Neural Network; RF, Random Forest; SVM, Support Vector Machine; NB, Naive Bayes; AdaBoost, Adaptive Boosting; XGBoost, Extreme Gradient Boosting; K-Fold, K-Fold cross validation; SHAP, Shapley Additive explanations; WBC, White Blood Cell; Cr, Creatinine; UA, Uric Acid; HbA1c, Glycosylated Hemoglobin; HDLC, High-density Lipoprotein Cholesterol.

### Data preprocessing

To reduce computational complexity and improve the credibility of the model, 50 out of 198 features were screened, according to medical literature and the clinical experience of physicians. These features included patient’s age, weight, WBC, blood lipid, Cr, etc. Some commonly used medical indicators are calculated in following formulae:

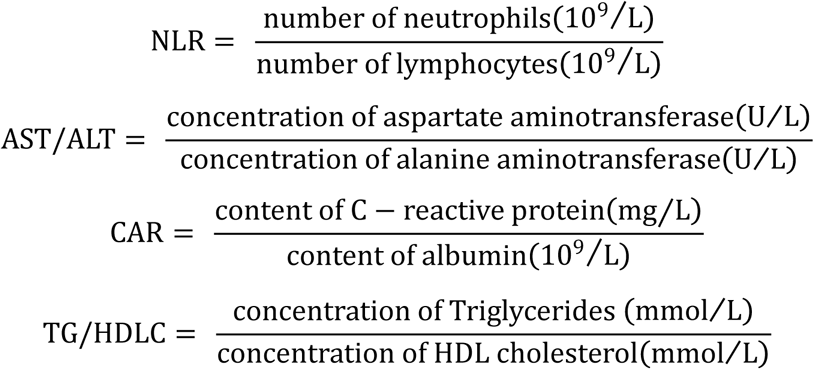

Abbreviations: NLR, neutrophil-to-lymphocyte ratio; AST, Aspartate aminotransferase; ALT, Alanine transaminase; CAR, ratio of C-reactive protein to albumin.

We utilized a beeswarm graph from the SHapley Additive exPlanation (SHAP) package to illustrate the final relationships between the hypertension and features collected so that we can identify whether features are risk factors or protective factors[13]; Consequently, some features were eliminated by combining the global feature importance graph and calculating the correlations. A comparison was made between the median imputation method, mean imputation method, and K-nearest neighbor algorithm methods for missing values imputation. Before initiating model training, outliers within the dataset were meticulously eliminated according to the 3-sigma principle. This process served to mitigate the disruptive influence of outliers on the model, thus enhancing the precision of the experimental results.

Initially, the data distribution was verified to decide whether to use the Spearman correlation coefficient or the Pearson correlation coefficient method. Secondly, the *p*-values were calculated for each feature to assess their impact on hypertension. Covariates with a *p*-value < 0.05 were then incorporated into the machine learning model. Following this, the outcomes of median imputation, mean imputation, and K-nearest neighbor algorithms were compared. After consulting with clinical experts, it was concluded that median imputation aligned better with the clinical characteristics of TG/HDLC and HDLC, while other indicators were more appropriate for the K-nearest neighbor algorithm. Consequently, a combination of these two imputation methods was adopted to impute the missing values. Finally, anomaly detection based on the 3-sigma principle was used to remove outliers and improve the model’s effectiveness with the data. The mean (μ) and Standard Deviation (σ) were calculated, and μ−3σ and μ+3σ were determined as thresholds, which were used to compare the data in the dataset. Finally, the data > μ+3σ or < μ−3σ were treated as outliers and were deleted. After the data preprocessing, the study population included 3380 hypertensive patients and 3696 non-hypertensive patients.

### Model construction

In order to construct the predictive model, eight machine learning algorithms were employed, including LR, DT, ANN, RF, SVM, NB, AdaBoost, and XGBoost. Although LR has “regression” in its name, it is more widely used for classification tasks. It is an extension of LR to estimate the probability of an event occurring. Decision Tree is a supervised learning algorithm that makes decisions through a tree structure. Artificial Neural Network is a computational model that emulates the structure and functionality of biological neural networks, enabling the estimation or approximation of complex functions that rely on the analysis of a vast quantity of input data. It is comprised of multitude of nodes (or neurons), which are usually distributed across distinct layers: input, hidden, and output layer. Random Forest is an algorithm based on a decision tree, which enhances the model’s generalisation ability and robustness by incorporating randomness into the training process. The Random Forest makes predictions by combining multiple decision trees, each trained on a randomly sampled dataset. The Support Vector Machine is one of the most popular machine learning methods. It is a supervised learning algorithm that can be used for classification or regression tasks. The fundamental concept of SVM is to identify an optimal hyperplane in the feature space that separates the samples of different classes and maximises the boundaries. Naive Bayes is a simple probabilistic classifier based on Bayes’ theorem, which assumes that features are independent of each other. In the training phase, it calculates the prior probability for each category and the conditional probability of each feature under each category. For a new instance, it calculates the posterior probability that the instance belongs to each category and then classifies the instance into the category with the highest posterior probability. Adaptive Boosting is an integrated learning algorithm that creates a strong classifier using a weighted combination of multiple weak classifiers. The weak classifiers are trained sequentially during the training process, with each classifier attempting to correct the errors of the previous classifier. Extreme Gradient Boosting is an optimised distributed gradient enhancement library. It is a fast, scalable, high-performance machine learning algorithm that can optimize model performance by adjusting several parameters, such as the eta, n_estimators, max_depth, min_child_weight, and regularisation. In addition, it can train decision tree models over numerous iterations and combine them to enhance prediction accuracy. As a high-performance machine learning algorithm, it has been widely recognised for its use in several areas. All models were constructed using the scikit-learn package in Python 3.10.13 (available at http://www.python.org, accessed on 22 May 2024). Grid Search was used to identify the optimal parameters for XGBoost.

### Model evaluation

The evaluation of models is a crucial aspect of understanding their classification capabilities. Currently, machine learning models for classification tasks are evaluated using several well-established metrics, including accuracy, precision, recall, F1-score and area under the curve (AUC), which are calculated as follows:

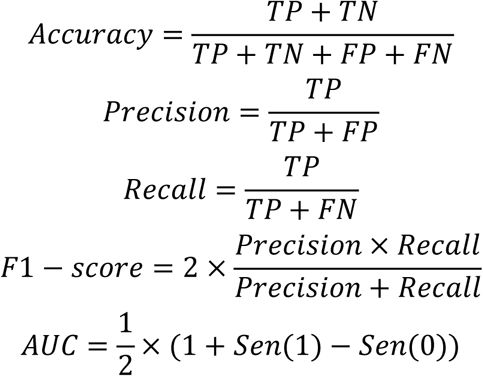

True positive (TP) is the number of correctly classified hypertensive patients. False positive (FP) denotes the number of healthy subjects who were predicted as hypertensive patients. True negative (TN) represents the number of correctly classified healthy subjects. False negative (FN) is the number of hypertensive patients who were classified as healthy individuals. Sen(1) represents the true positive case rate for the positive class of samples,while, Sen(0) denotes the true negative case rate for the negative class of samples. All the aforementioned metrics range from 0 to 1.

In addition, SHAP was employed to visualise model performance. SHAP interpretation originates from cooperative game theory and has a robust theoretical foundation. The method is a model-independent solution for model interpretability. After selecting the model with the best performance, SHAP was employed to quantify the marginal contribution of features to explain the model output and identify the salient features in different classifications. By assessing their positive or negative correlations, SHAP was able to discern whether an indicator serves as a risk factor or a protective factor for hypertension. Finally, beeswarm diagrams within the SHAP open-source software package were utilized to identify the associations between each indicator and hypertension.

The data preprocessing, model construction and evaluation were performed using Python Software version 3.10.13.

## Results

### Feature selection

The 50 variables included in this study were all continuous variables. Spearman’s correlation coefficient was calculated to analyse the correlation in order to further screen for features.

As illustrated in Table 1, age, weight, NLR, TG/HDLC, hypersensitive C-reactive protein (hs-CRP), white blood cell (WBC), platelets, total bilirubin, globulin, creatinine (Cr), uric acid (UA), triglyceride (TG), total bilirubin, high-density lipoprotein cholesterol (HDLC), glycosylated hemoglobin (HbA1c), fibrinogen, and prealbumin were all statistically significant (*P* < 0.05).

**Table 1.** Correlation analysis results for hypertension selection variables.

| Features | R-value | P-value |
| --- | --- | --- |
| Age | 0.452315561 | **4.52E-227** |
| Weight | 0.025567607 | **0.02** |
| NLR | 0.154255004 | **1.72E-25** |
| TG/HDLC | 0.032429651 | **0.02** |
| hypersensitive C-reactive protein | 0.029606069 | **0.04** |
| White blood cell | 0.075920922 | **3.18E-07** |
| Platelets | 0.046658731 | **0.00** |
| Neutrophil percentage | 0.16153046 | **7.97E-28** |
| Lymphocytes percentage | 0.15997334 | **2.57E-27** |
| Eosinophil percentage | -0.033448191 | **0.02** |
| Basophil percentage | ⁻0.033584537 | **0.02** |
| Aspartate aminotransferase | 0.051689599 | **0.00** |
| Alanine aminotransferase | 0.017739799 | 0.23 |
| Total bilirubin | 0.075111845 | **4.24E-07** |
| Globulin | 0.075900007 | **3.20E-07** |
| Albumin-globulin ratio | ⁻0.160788583 | **1.39E-27** |
| Alkaline phosphatase | 0.097178147 | **5.75E-11** |
| γ -glutamyltransferase | 0.02291171 | 0.12 |
| Total bilirubin | ⁻0.060773841 | **4.31E-05** |
| Indirect bilirubin | ⁻0.058804715 | **7.57E-05** |
| Creatine kinase | 0.035504106 | **0.01** |
| Urea | 0.102900232 | **3.99E-12** |
| Creatinine | 0.009328876 | **0.00** |
| Chlorion | 0.029371834 | **0.04** |
| Calcium | 0.019686893 | **0.01** |
| Uric acid | 0.023477643 | **0.01** |
| Magnesium | 0.005583856 | 0.70 |
| Diastase | 0.003350478 | 0.82 |
| Cystatin c | 0.307013667 | **2.37E-99** |
| Triglyceride | 0.052412456 | **0.00** |
| High density lipoprotein cholesterol | ⁻0.025622697 | **0.00** |
| Glycosylated hemoglobin | 0.330076618 | **1.92E-115** |
| Thyrotropin | 0.033452876 | **0.02** |
| Free triiodothyronine | -0.200776396 | **2.32E-42** |
| Prothrombin time | 0.002272608 | 0.87 |
| International standardization rate | 0.008134263 | 0.58 |
| Fibrinogen | 0.174097173 | **4.05E-32** |
| D - Dimer | 0.140401604 | **2.36E-21** |
| Prealbumin | 0.132183175 | **2.92E-13** |

NLR, neutrophil-to-lymphocyte ratio; TG/HDL, triglyceride/high density lipoprotein cholesterol; $P < 0.05$
is indicated in bold

In addition, the global importance feature map of SHAP was employed to identify additional features with the objective of optimising the final model performance. The top 20 most important features were included in the final model training (Fig. 2).

**Fig. 2.**
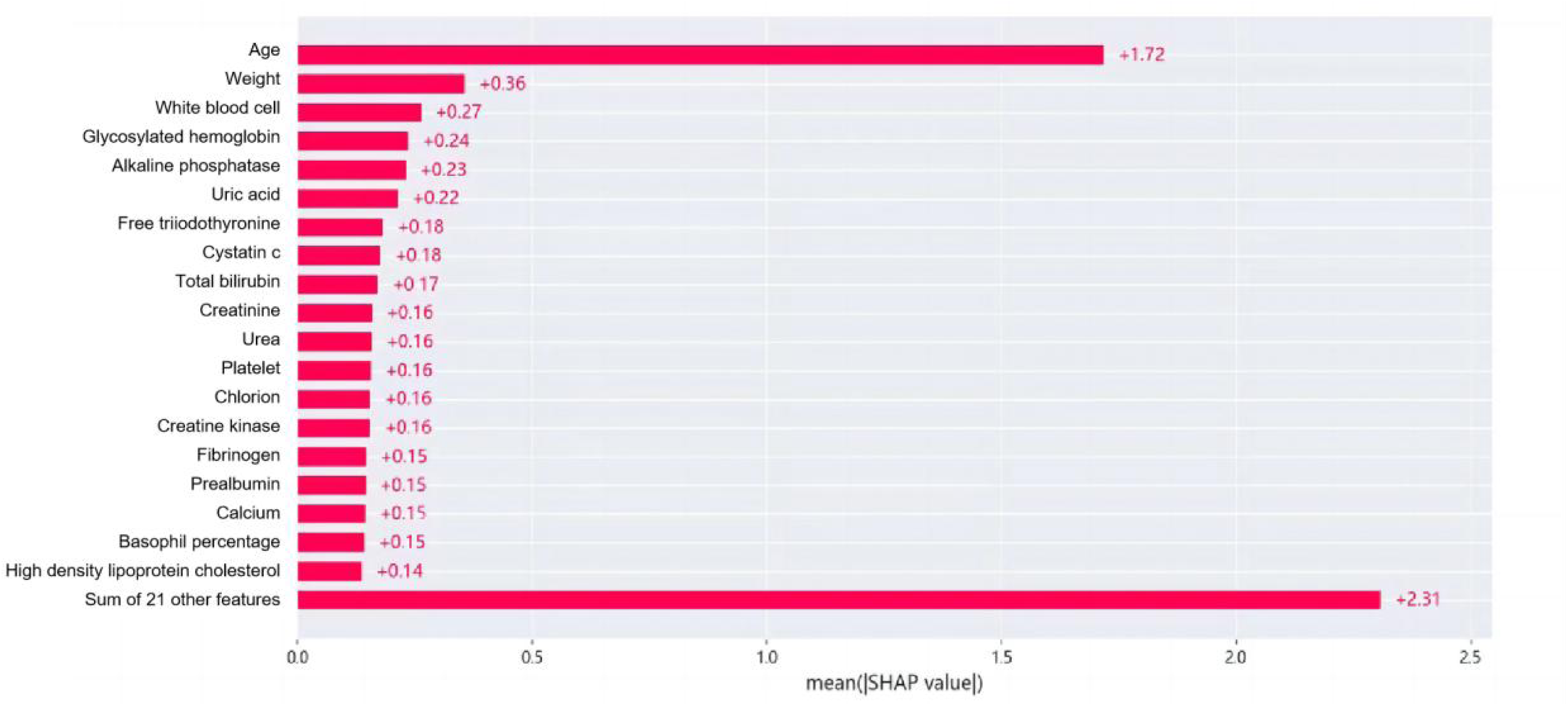
Top 20 important features.

### Data preprocessing

After data standardization and imputation of missing values, the data were subjected to anomaly detection based on the 3-sigma principle. Prior to data cleaning, The data volume was approximately balanced, and following data cleaning, and the changes in the data volume are presented in Table 2.

**Table 2.** Comparison of the sample size before and after data cleaning.

|  | HP | NHP | Total |
| --- | --- | --- | --- |
| Before sampling | 3363 | 3661 | 7024 |
| After sampling | 2903 | 3090 | 5693 |

HP, Hypertension; NHP, Non-hypertension

### Modeling and evaluation

The results of evaluation of the eight machine learning algorithms are presented in Table 3. The results demostrated that XGBoost exhibited the highest accuracy (0.7576 ± 0.0119), precision (0.7216 ± 0.0169), recall (0.7660 ± 0.0105), and F1-score (0.7430 ± 0.0104), outperforming the other models on all of these evaluation metrics.

**Table 3.** Comparison of the classification results of different models (mean ± SD)

|  | *Accuracy* | *Precision* | *Recall* | *F1-score* |
| --- | --- | --- | --- | --- |
| LR | 0.7093 ± 0.0081 | 0.6846 ± 0.0119 | 0.6761 ± 0.0142 | 0.6802 ± 0.0083 |
| DT | 0.6629 ± 0.0112 | 0.6305 ± 0.0148 | 0.6362 ± 0.0123 | 0.6332 ± 0.0097 |
| ANN | 0.7028 ± 0.0109 | 0.6569 ± 0.0126 | 0.7334 ± 0.0154 | 0.6929 ± 0.0104 |
| SVM | $0.7235 \pm 0.0141$ | $0.6774 \pm 0.0163$ | $0.7557 \pm 0.0123$ | $0.7143 \pm 0.0124$ |
| NB | $0.6845 \pm 0.0073$ | $0.6563 \pm 0.0067$ | $0.6508 \pm 0.0133$ | $0.6535 \pm 0.0097$ |
| AdaBoost | $0.7061 \pm 0.0113$ | $0.6701 \pm 0.0145$ | $0.7046 \pm 0.0093$ | $0.6868 \pm 0.0099$ |
| RF | $0.7522 \pm 0.0174$ | $0.7166 \pm 0.0236$ | $0.7591 \pm 0.0204$ | $0.7370 \pm 0.0166$ |
| XGBoost | **$0.7576 \pm 0.0119$** | **$0.7216 \pm 0.0169$** | **$0.7660 \pm 0.0105$** | **$0.7430 \pm 0.0104$** |

A confusion matrix is a formalised method for evaluating machine learning models, reflecting the results presented in Table 3. As can be easily deduced from Fig. 3, the overall performances of the RF and XGBoost models were significantly superior to those of the other models, with the XGBoost model having a slight edge over the RF model. Specifically, the XGBoost model slightly outperformed the RF in terms of accuracy, precision, and recall.

**Fig. 3.**
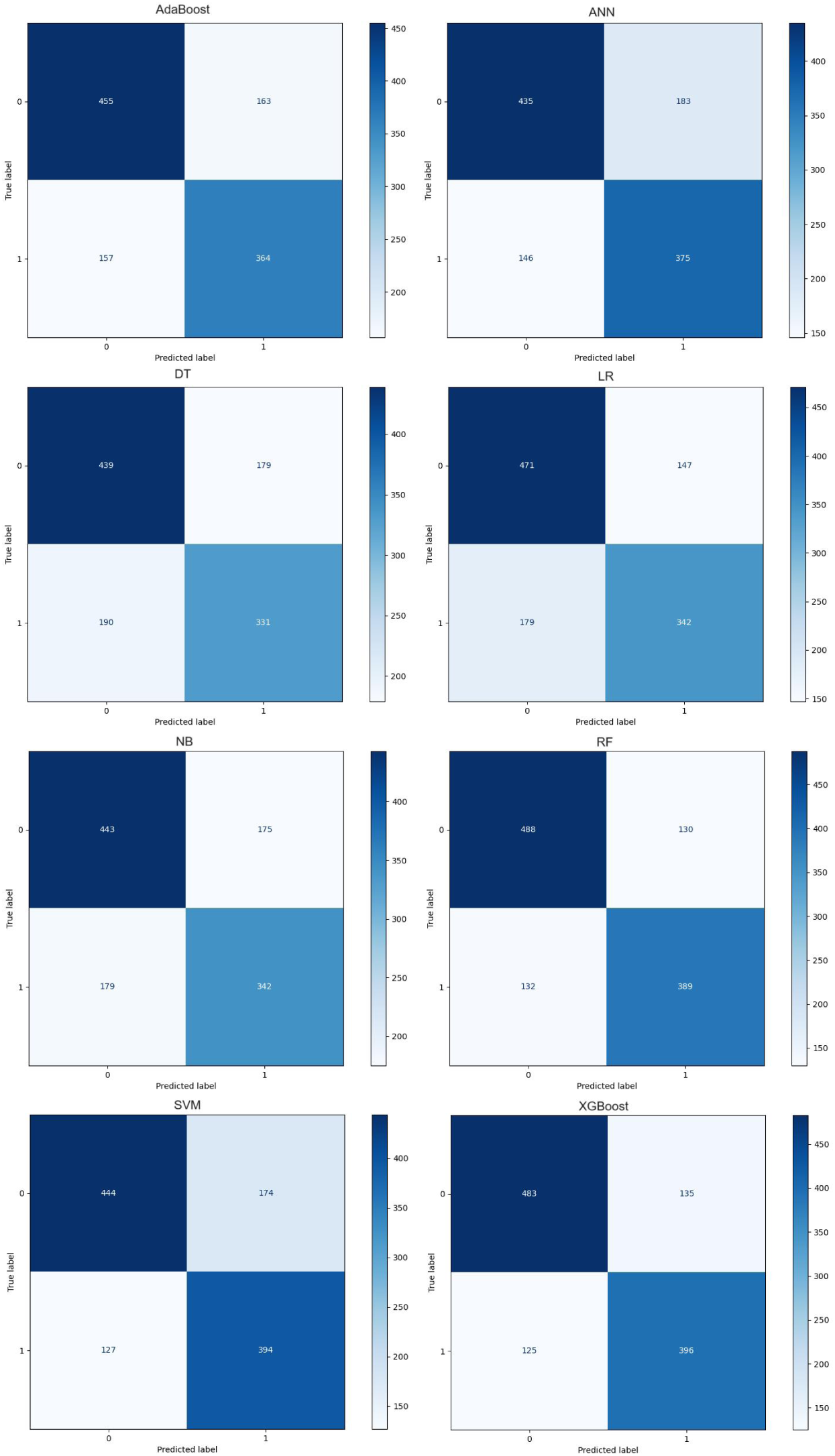
Model classification confusion matrix. LR, Logistic Regression; DT, Decision Tree; ANN, Artificial Neural Network; SVM, Support Vector Machine; NB, Naive Bayes; RF, Random Forest; AdaBoost, Adaptive Boosting; XGBoost, Extreme Gradient Boosting

The performance of the eight machine learning algorithms in predicting hypertension is presented in Fig. 4. While RF achieved the highest AUC of 0.847, XGBoost exhibited the closest performance with an AUC of 0.844. In contrast, DT exhibited the lowest AUC of 0.662.

**Fig. 4.**
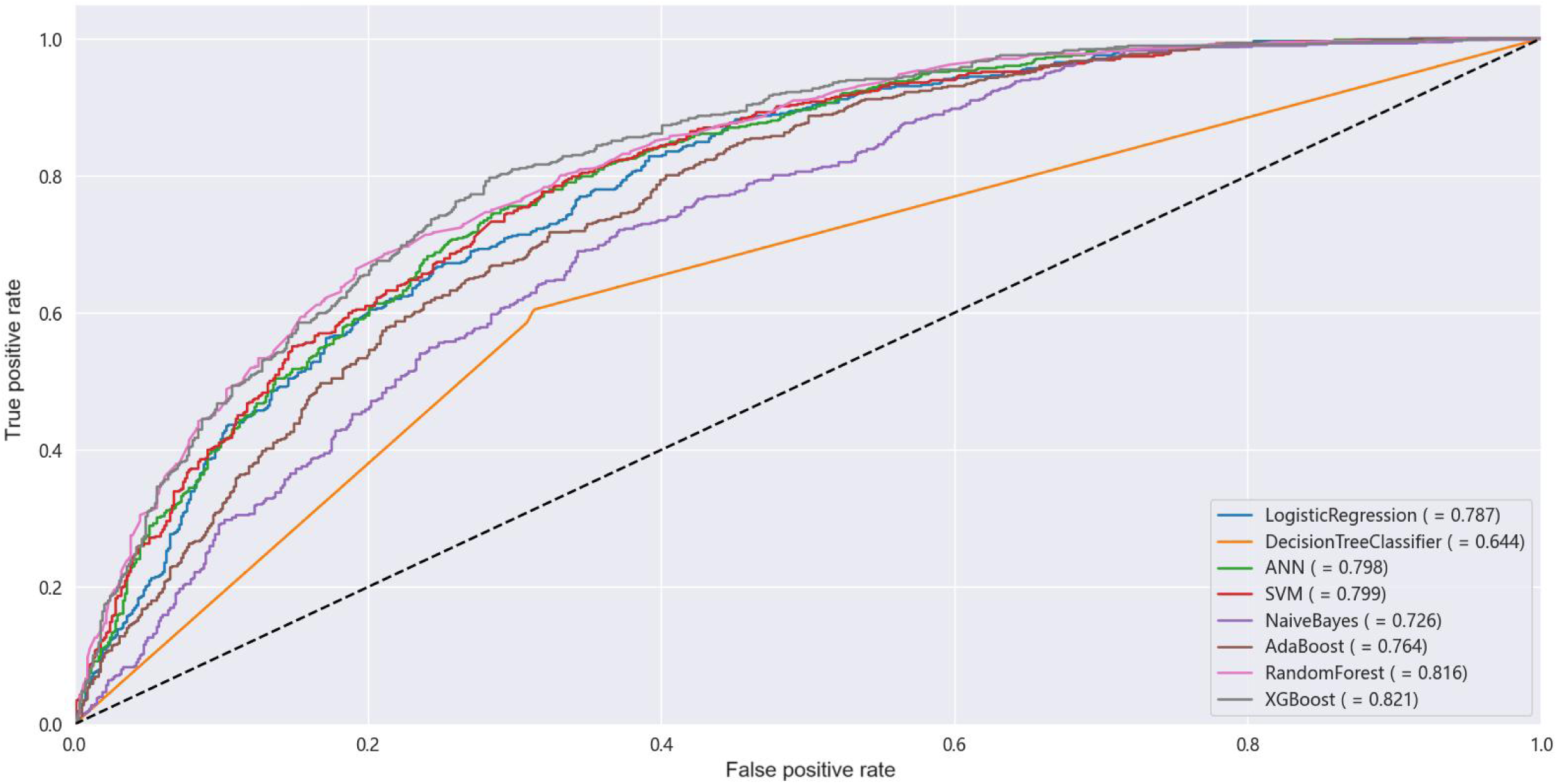
Receiver operating characteristic (ROC) curves for different classification models. ANN, Artificial Neural Network; SVM, Support Vector Machine; AdaBoost, Adaptive Boosting; XGBoost, Extreme Gradient Boosting

A comprehensive performance analysis of the eight models indicated that XGBoost was the optimal model for predicting hypertension. SHAP was employed to elucidate the relationship between the features and the output of the XGBoost model. As illustrated in Fig. 5, the top 10 indicators with the greatest impact on classification all exhibited a *P* value < 0.05. Fig. 5 illustrates that age, weight, WBC, Cr, UA, HbA1c, HDLC, and platelets were crucial features for hypertension prediction by XGBoost and significantly influencing the classification results. Age, weight, WBC, Cr, UA, HbA1c, and platelets were identified as risk factors, while HDLC was identified as a protective factor.

**Fig. 5.**
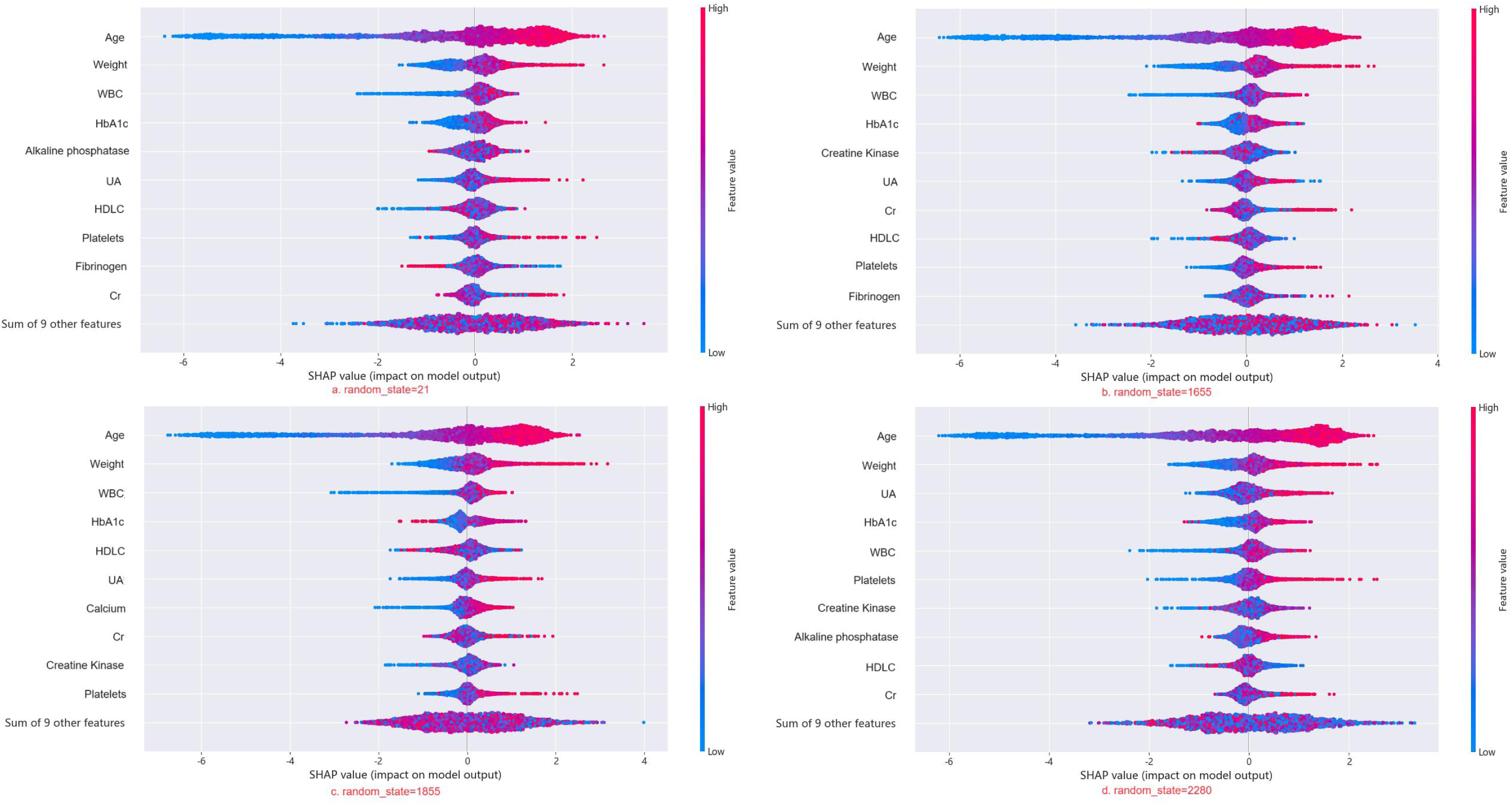
Comparison of XGBoost Model Interpretations using SHAP across Different Dataset Splits. SHAP, SHapley Additive exPlanation; WBC, White Blood Cell; Cr, Creatinine; UA, Uric Acid; HbA1c, Glycosylated Hemoglobin; HDLC, High-density Lipoprotein Cholesterol

## DISCUSSION

Hypertension is a prevalent adverse health outcome triggered by sleep disorders [14], and episodes of both can form a vicious cycle, increasing the risk of cardiovascular mortality [15]. Nevertheless, the correlation between clinical sleep disorders and the development of hypertension is frequently overlooked. Therefore, it is crucial to elucidate the clinical indicators associated with sleep disorders and hypertension in order to identify predictors of their occurrence. In this study, we constructed an XGBoost-based hypertension prediction model for individuals with sleep disorders using machine learning methods, and the higher recall of this model indicated its good reliability.

The developed model incorporates several features. Firstly, data from two centers in Nanjing, China were utilised for the analysis, with the patients’ clinical data subjected to standardised laboratory operations to guarantee the accuracy of the variables in the prediction model. Secondly, The presence of missing values and outliers is a common underlying problem in data processing. In this study, three methods were employed to address the issue of missing values: median, mean, and K-nearest neighbour interpolation. Outliers were handled using the 3-sigma principle, with the objective of ensuring the accuracy of the study, improving the quality of the data, and accelerating the speed of convergence of the model through strict data processing. Thirdly, eight different machine learning models were constructed, with the XGBoost model, demonstrating the highest accuracy, recall, F1 value, and precision. Finally, the SHAP method was employed to elucidate the relationship between the input features and output variables of the XGBoost model. The features were ranked according to their contribution to the model output, identifying several important hypertension-related indicators. This process enabled the objective extraction of key factors of disease occurrence from real clinical data.

Previous studies have attempted to develop predictive models for hypertension associated with sleep disorders. Khamsai et al. [16] used one-way logistic regression analyses to explore the risk factors for the development of hypertension in sleep disorders, but did not exclude the confounding effects of the risk factors on each other, which might have made the conclusions less accurate in larger samples. Shi et al. [17] employed machine learning to construct a model; however, such single-center sample size studies have the limitation of poor representativeness. Li et al. [18] collected multicenter data, but only selected a single model construction method, and the accuracy of the model was questionable. In addition, the characterisation data from these studies require specialized equipment, such as polysomnography, for monitoring purposes, which is costly to study and unsuitable for clinical dissemination. Consequently, the utilisation of multicenter datasets with readily available, larger, and well-structured data to construct a superior model of hypertension is warranted. Hypertension risk factors associated with sleep disorders identified in previous studies included age and body mass index [19,20]. In addition, the following novel findings remain from our study.

Elevated WBCs are a risk factor for hypertension in individuals with sleep disorders. Chronic sleep disorders induce WBC inflammatory gene transcription through activated partial inflammatory factor signalling, signal transducer and activator of transcription family proteins, and the nuclear factor kappa B (NF-κB) inflammatory signalling system, resulting an abnormal increase in WBC counts [21]. Fragmented sleep in sleep disorders also inhibits the secretion of orexin, which increases the secretion of colony-stimulating factor-1 from bone marrow cells, prompting the conversion of more bone marrow stem cells into WBCs [22]. Increased WBCs reduce vascular compliance by mediating vascular remodeling and affect vascular endothelial cell function to trigger vasodilation and contraction disorders, which in turn lead to the development of hypertension. WBCs adhere to the vascular endothelium and subendothelium through vascular and cell adhesion molecules [23]. This process causes vascular remodeling through the secretion of inflammatory factors that trigger the activation of matrix metalloproteinases, resulting in collagen deposition [24], which leads to the development of hypertension. In addition, pattern recognition receptors of WBCs during the inflammatory response cause irreversible vascular remodeling by affecting the proliferation and phenotypic transformation of vascular smooth muscle cells (VSMCs). This results in an increase in the thickness of the vascular wall and a decrease in the contractility of the VSMCs, also causing irreversible vascular remodeling [25]. Alternatively, nitric oxide (NO) produced by the endothelial nitric oxide synthase (eNOS) pathway is an important factor that mediates endothelium-dependent vascular relaxation [26]. Increased numbers of leukocytes reduce NO production through increased eNOS uncoupling, and desensitisation of the NO-Nitric oxide sensitive guanylyl cyclase (NOsGC)-cGMP pathway, which together trigger endothelial cell dysfunction [27]. In addition, sleep disorders catalyze reactive oxygen species (ROS) production through the activation of inflammatory factors that promote leukocytes secretion of myeloperoxidase and upregulation of NADPH oxidase activity in endothelial cells (ECs). Reactive oxygen species contribute to the uncoupling of eNOS and inactivation of NO [27,28], which mediate endothelial dysfunction-associated impaired vasodilatation, which in turn causes elevated blood pressure. In addition, ROS regulate the expression of the angiotensin II type 1 receptor (AT1R) on the cell surface through mechanisms such as foveolar protein phosphorylation [29], increasing the vasoconstrictor effect of angiotensin II (Ang II) and inducing the development of hypertension. The present study further validated the importance of WBCs in the development of sleep disorder-induced hypertension; however, whether there is a specific subpopulation of WBCs associated with increased risk remains to be further investigated.

Serum creatinine is a predictive risk factor for hypertension in patients with sleep disorders. Creatinine is an important indicator of renal function, and its metabolism is mainly accomplished in the kidneys. Creatinine excretion decreases when renal function is impaired, in addition to the kidneys being an important part of blood pressure regulation. Clinical studies have shown that patients with sleep disorders have a higher risk of renal impairment [30], and the mechanism might be the abnormal activation of sympathetic nerves in patients with sleep disorders, which causes endocrine dysfunction of the renin-aldosterone-angiotensin system, leading to an increase in the secretion of AngII and aldosterone [31]. AngII is mainly exerted through the AngII/AT1R pathway, which is widely distributed in VSMCs, to exert vasoconstriction, causing the peripheral blood pressure to increase and decrease. The vasoconstrictive function of AngII increases peripheral vascular resistance, thus causing an increase in circulating blood pressure. At the same time, AngII alters renal hemodynamics by constricting the renal vasculature, resulting in renal filtration impairment and sodium and water filtration limitation, which causes an increase in blood pressure associated with an increase in blood volume [32]. In addition, the same increase in aldosterone increases blood volume by reabsorbing sodium and water via activation of distal renal tubular epithelial Na channels (ENaCs), and over-activated ENaCs cause an increase in the membrane stiffness of ECs, blunting their vasodilator function [33] and decreasing their ability to regulate blood pressure.

We observed that elevated UA is a risk factor. When renal filtration is impaired due to sleep disorders, UA accumulates and precipitates as urate crystals, which further aggravate renal impairment when deposited in the renal tubules; and urate crystals deposited in the arteries cause arterial calcification, which reduces vascular compliance [34]. Patients with sleep disorders also increase endogenous UA production by increasing nucleotide turnover through elevated catecholamines following sympathetic activation [35,36], and high UA levels impair endothelial function by decreasing NO production via reduced eNOS phosphorylation [34]. In addition, UA induces VSMCs proliferation by stimulating angiotensinogen production [37], which benefits vessel wall thickening and ultimately contributes to the development of hypertension.

Glycated hemoglobin (HbA1c) is also a risk factor. Disorders of glycolipid metabolism are important mechanisms in the development of hypertension. Patients with sleep disorders often have abnormalities in their nervous and endocrine systems, such as sympathetic nerves and the hypothalamic-pituitary-adrenal axis, which affect the secretion of appetite-regulating hormones and cortisol in the body, thereby reducing the insulin sensitivity of the tissues [38,39] A decrease in glucose utilization causes an increase in the levels of late-phase glycosylation end-products [40], which increase the extracellular matrix area through protein adhesion [41]. The connected collagen is less susceptible to hydrolytic turnover, leading to thickening and sclerosis of the vascular wall [42], which triggers hypertension by decreasing vascular compliance. On the one hand, elevated blood glucose increases ROS production through the NF-κB pathway, and on the other hand, reduces NO production associated with elevated levels of asymmetric dimethylarginine, resulting in persistent endothelial dysfunction [43]. Studies have shown that disorders of glucose and lipid metabolism are closely related to insulin resistance, and that reduced insulin sensitivity decreases the specific inhibition of triglyceride-rich very-low-density lipoprotein (VLDL) by insulin, while decreasing the activity of lipoprotein lipase, which breaks down VLDL, ultimately leading to elevated cholesterol-rich ester residues in the blood, causing secondary atherosclerosis [44], reduced elastic storage of large arteries, and reduced cushioning of blood pressure. In people with elevated blood glucose, the combined use of statins can effectively reduce the incidence of atherosclerotic cardiovascular disease [45], suggesting that we should be more thorough in considering the use of medications in people at high risk for sleep disorders.

High-density lipoprotein cholesterol (HDLC) prevents hypertension from occurring in sleep disorders. Clinical studies have shown that sleep disorders are associated with low HDLC levels [46]. Increased secretion of inflammatory factors induced by sleep disorders inhibits apolipoprotein A-I (ApoA-I) expression, resulting in lower levels of HDLC produced by the liver. In addition, serum amyloid A (SAA), which is increased under inflammation, replaces inhibited ApoA-I in HDLC, and SAA-rich HDLC is susceptible to rapid clearance [47]. This ultimately leads to a decrease in the total amount of HDLC. Normal levels of HDLC bind to scavenger receptor BI on the endothelium, stimulating the activation of eNOS and the release of more NO [48], reducing the activity of NADPH oxidase in ECs, and increasing the bioavailability of NO [49], which protects the function of ECs and reduces the risk of hypertension.

Among the outputs of this model, risk predictors that are closely associated with the chance of developing hypertension in patients with sleep disorders also included platelets, which in the case of vascular microinjury, can aggregate and adhere to the injury in large numbers, causing vascular sclerosis and triggering hypertension [50]. Alkaline phosphatase is also one of the risk factors, which is mainly used as an indicator of liver injury, thus the link with sleep disorders and hypertension requires further discussion. Fibrinogen and creatine kinase are more confounding in the output and their predictive tendencies could not be accurately identified.

In this model, the top ten predictors strongly associated with the development of hypertension are age, weight, WBC, UA, Cr, HbA1c, HDLC, platelets, alkaline phosphatase, and creatine kinase, which respond to the effects of sleep disorders on multiple systems of the body and emphasize the need for early personalized management and treatment of sleep disorders to prevent the further development of hypertension.

Our study has some limitations. First, because of the high volatility of some data, certain features are not well recognized in our model results, such as fibrinogen and creatine kinase; however, they do not affect the accuracy of the model, and we will further improve the prediction model through data collection and algorithm optimization. Second, most of the indexes that we included in the model are blood-related parameters, and to explore the effects of sleep disorders on hypertension more comprehensively, future studies will require different types of examination parameters. Finally, the sleep disorder population we included was limited to Nanjing, which might lack generalisability to a wider area.

## Conclusion

Our study established the best risk prediction model for hypertension in patients with sleep disorders using a large amount of commonly collected clinical data, which provides a reference for clinical interventions to reduce the prevalence of hypertension, and will help doctors to further judge treatment plans and prognosis.

### Perspectives

1. In recent years, the incidence of sleep disorders has increased, and the number of people with hypertension caused by sleep disorders has also begun to increase. Restricted by the complexity of the patient’s medical records and limited medical resources in the clinic, these patients with hypertension are usually diagnosed and then traced back to the cause of the disease through specialized examination, but at this time, the adverse consequences of hypertension have already been caused, and the patient needs to take medication to control their blood pressure for a long period of time. Our study closely monitors the etiology in the early stages and screens high-risk groups through risk factor control in common clinical blood examination, which is clinically important for preventing the occurrence of hypertension and reducing the healthcare burden.
2. Major hospitals under medical informatization are beginning to use electronic medical records to record structured information such as patients’ diagnoses, examinations, and laboratory tests, and machine learning can predict the trend of these complex data thus achieving the function of disease prediction and assisting clinical decision-making. In previous cross-sectional studies, there was a single risk factor for hypertension in sleep disorders, and the prediction results lacked stability due to sample and model selection. In our study, eight models were built and compared by machine learning, and the most stable and accurate model was finally selected to make the final results more clinically applicable.

### What Is New?

This study is the first to explore the risk factors for the development of hypertension in sleep disorders through common clinical indicators, and multiple models were used to ensure the accuracy and interpretability of the final results.

### What Is Relevant?

Ten factors such as age, weight, white blood cell, uric acid, creatinine, and other factors that are strongly associated with the development of hypertension in sleep-disordered populations were identified through machine learning.

### Clinical/Pathophysiological Implications?

The risk factors identified in this study can be used as serum markers for the development of hypertension in people with sleep disorders, which can help physicians to quickly and easily identify high-risk groups and take relevant interventions in their daily practice.

## Data Availability

The data for this study are obtained from the electronic medical systems of Jiangsu Provincial Hospital of Chinese Medicine and Jiangsu Provincial Government Hospital. For patient privacy protection, the data of this study are only provided to some of the collaborators. Data supporting the study results can be obtained from the authors upon reasonable request.

## Nonstandard Abbreviations and Acronyms

LR: Logistic Regression
DT: Decision Tree
ANN: Artificial Neural Network
RF: Random Forest
SVM: Support Vector Machine
NB: Naive Bayes
XGBoost: Extreme Gradient Boosting
AdaBoost: Adaptive Boosting
AUC: area under the curve
SHAP: SHapley Additive exPlanation
WBC: white blood cell count
Cr: creatinine
UA: uric acid
HbA1c: glycosylated hemoglobin
HDLC: high-density lipoprotein cholesterol
NLR: neutrophil-to-lymphocyte ratio
AST: Aspartate aminotransferase
ALT: Alanine transaminase
CAR: ratio of C-reactive protein to albumin
TG: triglyceride
hs-CRP: hypersensitive C-reactive protein
NF-κB: nuclear factor kappa B
VSMCs: vascular smooth muscle cells
NO: nitric oxide
eNOS: endothelial nitric oxide synthase
NOsGC: NO-Nitric oxide sensitive guanylyl cyclase
ROS: reactive oxygen species
ECs: endothelial cells
AT1R: angiotensin II type 1 receptor
Ang II: angiotensin II
ENaCs: epithelial Na channels
VLDL: very-low-density lipoprotein
ApoA-I: apolipoprotein A-I
SAA: serum amyloid A

## Acknowledgments

We are grateful to all the authors and participants who were involved in organizing the data and writing the code.

## Sources of Funding

This work was supported by the grants from the National Natural Science Foundation of China (grant number 82274631), the Key Research and Development Plan of Jiangsu Province (Social Development, BE2021751), the Key Research and Development Plan of Jiangsu Province (Social Development, BE2023793) and Jiangsu Commission of Health (Prevention Subject, Ym2023105).

## Disclosures

None.

## Data Availability Statement

Data from this study cannot be shared due to privacy terms and are available from the authors upon reasonable request. Dr. Liu has full access to all the data in this study and takes responsibility for its integrity and the data analysis.

## Notes

### Competing Interest Statement

The authors have declared no competing interest.

### Clinical Trial

Registration number: ChiCTR2200059161.

### Author Declarations

The Ethics Committee of Jiangsu Provincial Hospital of Chinese Medicine (Approved No. of ethic committee: 2021NL-202-02).

